# Effect of Vesicular Monoamine Transporter Type-2 Inhibitors on Motor Features in HD

**DOI:** 10.64898/2026.09.26.26364085

**Authors:** Nabil Halabi, Annie Killoran, Peg C. Nopoulos

## Abstract

**Background:** Huntington’s disease (HD) is an inherited neurodegenerative disorder primarily associated with motor and psychiatric manifestations. No disease-modifying treatment currently exists, with vesicular monoamine transporter type-2 inhibitors (VMAT2i’s) being the only FDA-approved therapy for motor features in HD. Previous studies evaluated the antichorea effects of VMAT2i’s but did not address its effects on non-choreiform features.

**Objectives:** This study aimed to characterize the effect of VMAT2i’s on hyperkinetic, hypokinetic and total hypokinetic subscores of the UHDRS motor assessment in patients with motor-manifest HD from the *Enroll-HD* research platform.

**Methods:** Data from 622 participants were analyzed. Linear mixed effects models were constructed to compare the rates of change of choreiform and non-choreiform motor scores before and after the initiation of a VMAT2i. We calculated and plotted the direction, magnitude and significance of VMAT2i administration on the rates of change of motor subscores. A similar analysis was performed investigating the effect of antipsychotic medications on motor subscales in HD.

**Results:** VMAT2i’s reversed progression of hyperkinesis but significantly increased the rates of change in the hypokinetic and total hypokinetic subscores. Antipsychotics also displayed efficacy in reversing progression of chorea but did not significantly influence progression of hypokinesis.

**Conclusions:** The initiation of VMAT2i’s was associated with an improvement in hyperkinesis but was associated with worsening of other motor features of HD. Hypokinesis increases progressively in HD and is correlated with functional impairment and global decline. Therefore, the use of VMAT2i’s to treat motor features in HD should be a nuanced decision, considering its multipronged impact.

## 1 Introduction

Huntington’s disease (HD) is a hereditary neurogenerative disease caused by a cytosine-adenine-guanine (CAG) trinucleotide repeat expansion in the huntingtin (*HTT*) gene, located on the short arm of chromosome 4.(1) HD is known for its progressive symptoms leading to significant disability and mortality.(1) Much of the morbidity is derived from the complex symptomatology of HD, which includes motor, psychiatric, and cognitive manifestations.(1) Moreover, there are currently no disease-modifying therapies for HD, which has a 100% fatality rate in all those who inherit the mutation.(1) Until a curative intervention is discovered, supportive treatment is the mainstay of HD management.(2) As such, the primary goals of care-providers are to: ameliorate symptoms, promote functionality/independence and palliate throughout the disease process.(3)

To achieve these aims, it is essential to optimize management of the motor features of HD. HD is classically identified by choreiform movements.(4) This is largely true of the initial phase of the disease, where hyperkinesis first manifests.(5) As the disease progresses, chorea typically diminishes in severity, giving way to hypokinesis.(5, 6) This classically includes bradykinesia, rigidity, postural instability, and gait disturbance.(7)

The FDA has only approved three drugs for the treatment of HD: tetrabenazine, deutetrabenazine and valbenazine.(8, 9)These medications all belong to the same class, namely vesicular monoamine transporter type-2 inhibitors (*i.e.* VMAT2i’s). VMAT is a cytosolic transporter protein bound to the membranes of secretory vesicles located in the axon terminals of presynaptic, monoaminergic (*i.e.* serotonergic, histaminergic, noradrenergic, and/or dopaminergic) neurons.(10) VMAT2i’s function by inhibiting this enzyme and hence monoaminergic neurotransmission. VMAT2i’s are specifically indicated for the treatment of chorea in Huntington’s disease.(11)

Despite not constituting curative treatment, VMAT2i’s remain the mainstay of medical management in HD.(1) New evidence has called into question the use of VMAT2i’s for managing all of HD’s motor features. Dopamine depletion can attenuate the hyperkinetic movements of HD, while accentuating late-stage hypokinesis.(7) However, a previous analysis from this group recently showed that *most* motor features in HD progress along a hypokinetic-like trajectory.(12) Therefore, HD may be best described as a mixed movement disorder.

Furthermore, evidence suggests that HD’s hypokinetic features worsen with increasing age and CAG length,(7, 13) and are strongly correlated with worsening cognitive performance,(14) lower functional status,(15, 16) and a poorer global condition.(15) A similar relationship between hyperkinetic features and cognitive measures has not been identified.(14)

Previously, the effect of tetrabenazine was limited to evaluating change in chorea scores as the primary outcome of experimental investigations.(9, 17) However, given that VMAT2i’s may worsen hypokinetic features, the current gap in our knowledge is the effect of VMAT2i’s on the entirety of the Unified Huntington’s Disease Rating Scale (UHDRS) motor subscales. Given our previous work which elucidated that the majority of the UHDRS motor features follow a hypokinetic trajectory(12), there is a need to investigate the effect of VMAT2i’s on all 31 sub-scales that make up the UHDRS Total Motor Score (*i.e.* TMS).

## 2 Methods

### 2.1. Study Design

The data utilized for secondary analysis in this paper is derived from the *Enroll-HD* research platform. Facilitated by the *Cure Huntington’s Disease Initiative* (CHDI), this platform provides large datasets to clinical researchers derived from their studies: *ENROLL-HD* (a multi-site, longitudinal study of HD patients and controls) and *REGISTRY* (a predecessor study). At each study visit, the UHDRS is administered to assess the participant’s motor status. The total number of study participants across the dataset was 25,550 at the time of analysis.

This investigation studied the effect of VMAT2i’s on the UHDRS motor items. Analyses were limited to gene-expanded (*i.e.* CAG > 35) participants who had a diagnostic confidence level of 4 (DCL-4); i.e. individuals with 99% diagnostic confidence in pronouncing HD.(18) Participants with missing CAG repeat lengths were excluded. Pharmacological information is also recorded at each participant’s visit, including medication start and stop dates. This makes it possible to study motor progression in the epochs before and after drug administration (i.e. VMAT2i). If a subject had multiple therapeutic trials, the earliest one was selected to avoid introducing the confounder of repeated drug exposures.

To avoid selection bias in comparing VMAT2i users to non-users, each participant served as their own control, mimicking a single-subject experimental design.(19) Specifically, we included VMAT2i users with at least three study visits. Participants had to have at least two visits where they were not taking a VMAT2i and at least one visit after initiation. In so doing, we obtain sufficient data to calculate the rate of progression of motor features *before* VMAT2i initiation and the rate of progression *after* VMAT2i initiation (*Supplementary Figure 1*). Additionally, for participants who had more than the minimum three-visit structure (*e.g.* those with several visits before / after VMAT2i initiation), all available visits that occurred after a motor diagnosis but before VMAT2i initiation were included. Furthermore, any visit where a subject was still taking a VMAT2i was included for analysis. Throughout the duration of *Enroll*, most follow-ups occurred within 1 year of the previous visit. Some inter-visit intervals are quite large, spanning several years. A maximum limit of 1.5 years was imposed as the largest acceptable inter-visit interval. Ultimately, these criteria allowed for the maximum number of participants to be selected, while still maintaining a valid temporal structure to infer the effect of VMAT2i use. Lastly, participants who had missing datapoints for any of the 31 UHDRS motor items were also excluded from analysis.

This analysis adapted an innovation from our previously mentioned work; namely, the use of composite motor scores based on shared trajectory.(12) We adapted the average hyperkinetic subscore (HyperS), which incorporates all seven choreiform measures from the UHDRS TMS: facial, buccal-oral-lingual (BOL), truncal, right upper extremity (RUE), left upper extremity (LUE), right lower extremity (RLE) and left lower extremity (LLE) chorea subscores. The original average hypokinetic subscore (HypoS) was composed of: bodily bradykinesia, right arm rigidity, left arm rigidity and retropulsion push test (i.e. classical hypokinetic features (7)). In this analysis, we added three more hypokinetic measures: truncal dystonia, tandem-walking and gait. These are all markers of postural instability, which is strongly associated with a hypokinetic phenotype as well.(20) Finally, we adapted the total hypokinetic subscore (THypoS) as is, constituting the 24 non-choreiform UHDRS motor subscales. The reason why all 24 non-choreiform subscales were classified as hypokinetic is due to their similarity in trajectory to the classical hypokinetic features, which seems to imply a shared underlying pathoetiology.(12)

### 2.2. Statistical Analysis

Statistical analysis was performed using the *R* programming language (version 4.2.1) in *Rstudio* 2022.07. Our primary analysis compared the rates of progression of the composite motor subscores previously described. Each individual motor subscale is scored from 0 to 4, separated by whole number increments, with 0 representing absence of the abnormality and 4 representing full severity. The HyperS and HypoS subscales each included 7 representative hyperkinetic or hypokinetic subscales and have a total possible score of 28 (i.e., 4 x 7). The THypoS, in contrast, has a total possible score of 96 (i.e., 4 x 24). The change in HyperS, HypoS, and THypoS was calculated between each two consecutive visits, denoted as Δ *HyperS*, Δ *HypoS* and Δ *THypoS*. Finally, an additional variable denoted *epoch* was added to each row of data, indicating whether the given row occurred before (*epoch* = 1) or after (*epoch* = 2) VMAT2i administration.

Thereafter, three linear mixed-effects models were constructed, comparing the mean change in HyperS, HypoS, and THypoS before (epoch 1) and after (epoch 2) VMAT2i initiation. Each model included CAG repeat length, age, sex (*i.e.* assigned gender at birth, AGAB), disease duration and HD Integrated Staging System (HD-ISS) stage as covariates. We also included the covariate of concomitant antipsychotic use to account for the potential antidopaminergic impact this medication class may contribute. A random effect term for each participant was also included, to account for intra-subject and inter-subject variability. For each model predicting HyperS, HypoS, and THypoS, the difference in the annualized change between the pre-VMAT2i and post-VMAT2i time periods was compared. For visualization purposes, these linear mixed effects models were repeated using the continuous variable *time in study*. In said models, the date of VMAT2i initiation was normalized to *time= 0 years*. Negative values represent time before VMAT2i initiation and positive values represent time thereafter. The continuous variable representing *time in study* was constructed as a non-linear value with a knot placed at time equals zero, to show how the trajectory of each motor score changed before and after initiation of a VMAT2i.

We performed secondary analyses to evaluate the effect of antipsychotics on HyperS, HypoS, and THypoS. These models were identical to those used for the primary analysis. However, instead of evaluating the periods before and after initiation of VMAT2i’s, we compared the periods before and after initiation of an antipsychotic. In these models, VMAT2i use was included as a covariate, analogous to antipsychotic use as a covariate in the primary models.

### 2.3. Data Sharing

The data utilized in this paper’s secondary analysis is derived from the *Enroll-HD* clinical research platform, provided by CHDI. It is available for qualified researchers worldwide who are interested in using this large dataset to further research on HD.

## Results

There were 622 participants eligible for primary analyses. *Figure 1* summarizes the details of the selection process. *Table 1* outlines the participants’ baseline characteristics. The average age was 51.6 ± 12.2 years and there were slightly more individuals assigned female at birth (AFAB) compared to those assigned male (AMAB) (324 vs. 298). The average age at clinical diagnosis was 47.8 ± 12.1 years, which is commensurate with HD’s natural history.(21) Approximately half (48.6%) of participants were concomitantly using an anti-psychotic. The average TMS was 36.2 ± 17.4, with average subscores of 10.2 ± 4.90 (HyperS), 6.57 ± 2.29 (HypoS) and 26.0 ± 14.6 (THypoS). Most participants were HD-ISS Stage three (54.2%), with an average disease duration of 3.81 ± 3.42 years and an average disease burden of 417 ± 87.6; all typical for a cohort of motor-manifest adult HD patients.(22) Disease burden (i.e. [CAG – 35.5] x Age) is a cumulative measure which represents the patient’s stage in the disease course.(23) The average total daily dose, at baseline, for the VMAT2i’s are as follows: 43.87 mg (tetrabenazine), 32.8 mg (deutetrabenazine), and 62.9 mg (valbenazine). The corresponding average total daily doses for the top three most-used antipsychotics were: 5.8 mg (olanzapine), 1.5 mg (risperidone), and 81 mg (quetiapine). The doses of VMAT2i’s are in the appropriate range for an antichorea effect(24), while the dosages of the antipsychotics are at the lower range of normal, suggesting that the indication was mood stabilization and not psychosis.(25)

**Figure 1:**
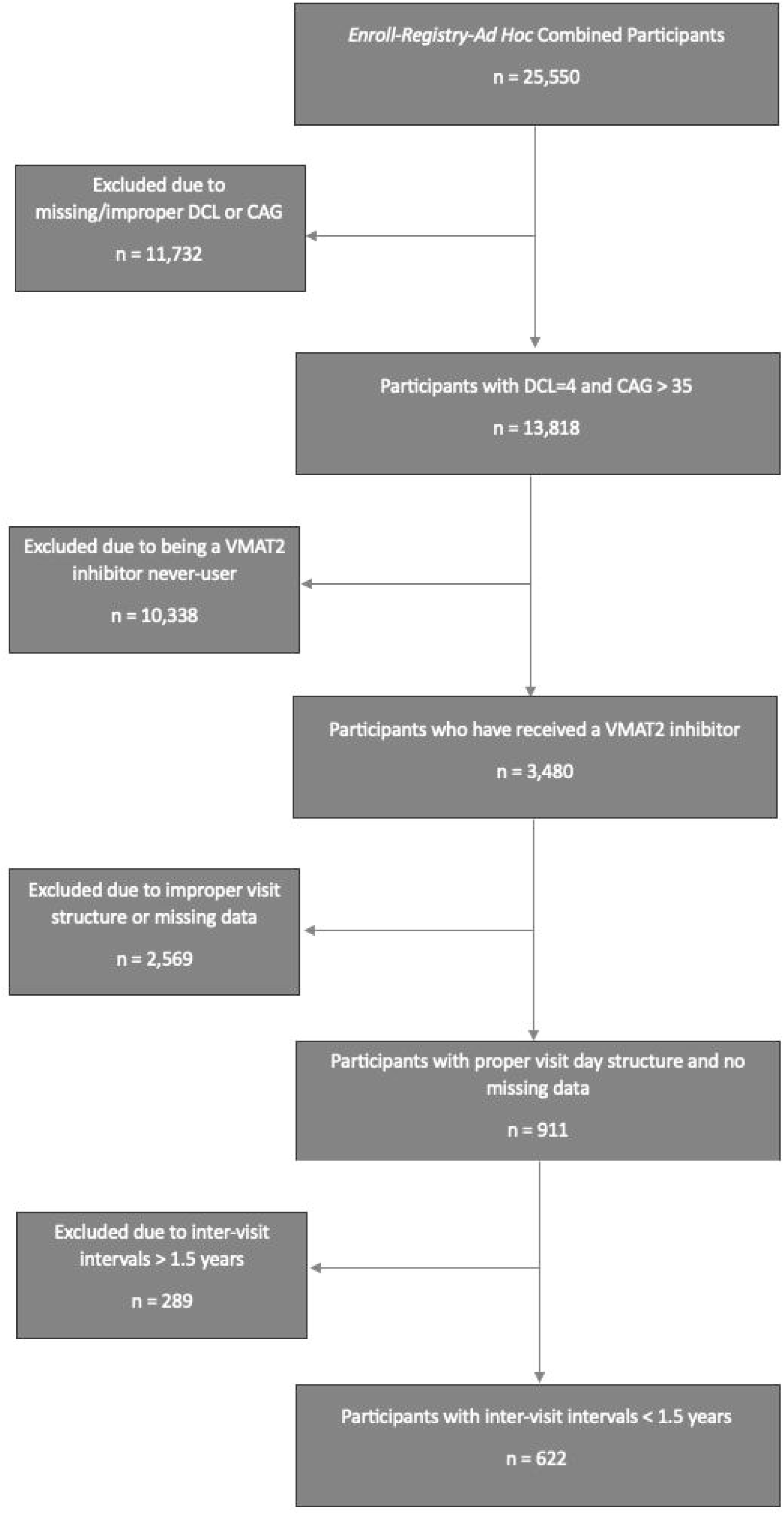
Flowchart of participant selection. DCL: diagnostic confidence level; CAG: cytosine-adenine-guanine; VMAT2; vesicular monoamine transporter type-2.

**Table 1:** Baseline demographics of VMAT2i users. Yrs: years; BMI: body mass index; kg: kilogram; m^2^: squared meter; SD: standard deviation; VMAT2i: vesicular monoamine transporter type-2 inhibitors; CAG: cytosine-adenine-guanine; min: minimum; max: maximum.

| **Demographic** | **Value** |
| --- | --- |
| **Age (yrs.)** |  |
| Mean (SD) | 51.6 (12.2) |
| Median [Min, Max] | 52.0 [21.0, 83.0] |
| **Assigned Gender at Birth** |  |
| Assigned Male at Birth (%) | 298 (47.9%) |
| Assigned Female at Birth (%) | 324 (52.1%) |
| **Calculated BMI (kg/m<sup>2</sup>)** |  |
| Mean (SD) | 24.7 (4.30) |
| Median [Min, Max] | 24.1 [15.4, 45.1] |
| Missing (%) | 28 (4.5%) |
| **Duration of Disease (yrs.)** |  |
| Mean (SD) | 3.81 (3.42) |
| Median [Min, Max] | 3.00 [-1.00, 18.0] |
| Missing | 10 (1.6%) |
| **Age at Clinical Diagnosis** |  |
| Mean (SD) | 47.8 (12.1) |
| Median [Min, Max] | 48.0 [17.0, 80.0] |
| Missing (%) | 10 (1.6%) |
| **Use of Antipsychotics** |  |
| Users (%) | 302 (48.6%) |
| Never-Users (%) | 320 (51.4%) |
| **Disease Burden** |  |
| Mean (SD) | 417 (87.6) |
| Median [Min, Max] | 417 [135, 969] |
| **Age of Death (yrs.)** |  |
| Mean (SD) | 58.7 (12.8) |
| Median [Min, Max] | 58.0 [29.0, 86.0] |
| Missing (%) | 515 (82.8%) |
| **Cause of Death** |  |
| Pneumonia (%) | 18 (2.9%) |
| Other Infection (%) | 7 (1.1%) |
| Cancer (%) | 4 (0.6%) |
| Stroke (%) | 1 (0.2%) |
| Trauma (%) | 2 (0.3%) |
| Suicide (%) | 6 (1.0%) |
| Other (%) | 30 (4.8%) |
| Missing (%) | 554 (89.1%) |
| **Total Motor Score (TMS)** |  |
| Mean (SD) | 36.2 (17.4) |
| Median [Min, Max] | 34.0 [4.00, 112] |
| **Hyperkinetic Subscore (HyperS)** |  |
| Mean (SD) | 10.2 (4.90) |
| Median [Min, Max] | 10.0 [0, 28.0] |
| **Hypokinetic Subscore (HypoS)** |  |
| Mean (SD) | 6.57 (4.29) |
| Median [Min, Max] | 6.00 [0, 25.0] |
| **Total Hypokinetic Subscore (THypoS)** |  |
| Mean (SD) | 26.0 (14.6) |
| Median [Min, Max] | 24.0 [0, 91.0] |
| **Ethnicity** |  |
| White (%) | 590 (94.9%) |
| Black (%) | 5 (0.8%) |
| Latinx (%) | 10 (2.4%) |
| Native American (%) | 1 (0.3%) |
| Mixed (%) | 2 (0.8%) |
| Asian (%) | 5 (0.6%) |
| Other (%) | 9 (1.5%) |
| **CAG Repeats** |  |
| Mean (SD) | 44.2 (3.59) |
| Median [Min, Max] | 44.0 [38.0, 66.0] |
| **HD-ISS Stage** |  |
| 0 | 0 (0%) |
| 1 | 1 (0.2%) |
| 2 | 38 (6.1%) |
| 3 | 337 (54.2%) |
| Missing | 246 (39.5%) |
| **Average VMAT2i Total Daily Dose (in milligrams, 3 significant figures)** |  |
| Tetrabenazine | 43.9 |
| Deutetrabenazine | 32.8 |
| Valbenazie | 62.9 |

Prior to initiation of a VMAT2i, the mean annualized change in HyperS was 0.97 (Standard Error 0.28) points per year (p.p.y.). After a VMAT2i was initiated, the mean annualized change became -0.86 p.p.y. (S.E. 0.28), which was significantly less (p<0.0001; *Figure 2A*). The mean annualized change in HypoS before VMAT2i initiation was 0.90 p.p.y. (S.E. 0.17). However, after a VMAT2i was initiated, the rate of change of HypoS worsened significantly to 1.27 p.p.y. (S.E. 0.18; p=0.031; *Figure 2B*). The mean annualized change in THypoS before VMAT2i initiation was 3.27 p.p.y. (S.E. 0.50). This worsened significantly after initiation to 4.24 p.p.y. (S.E. 0.51; p=0.0487; *Figure 2C*).

**Figure 2:**
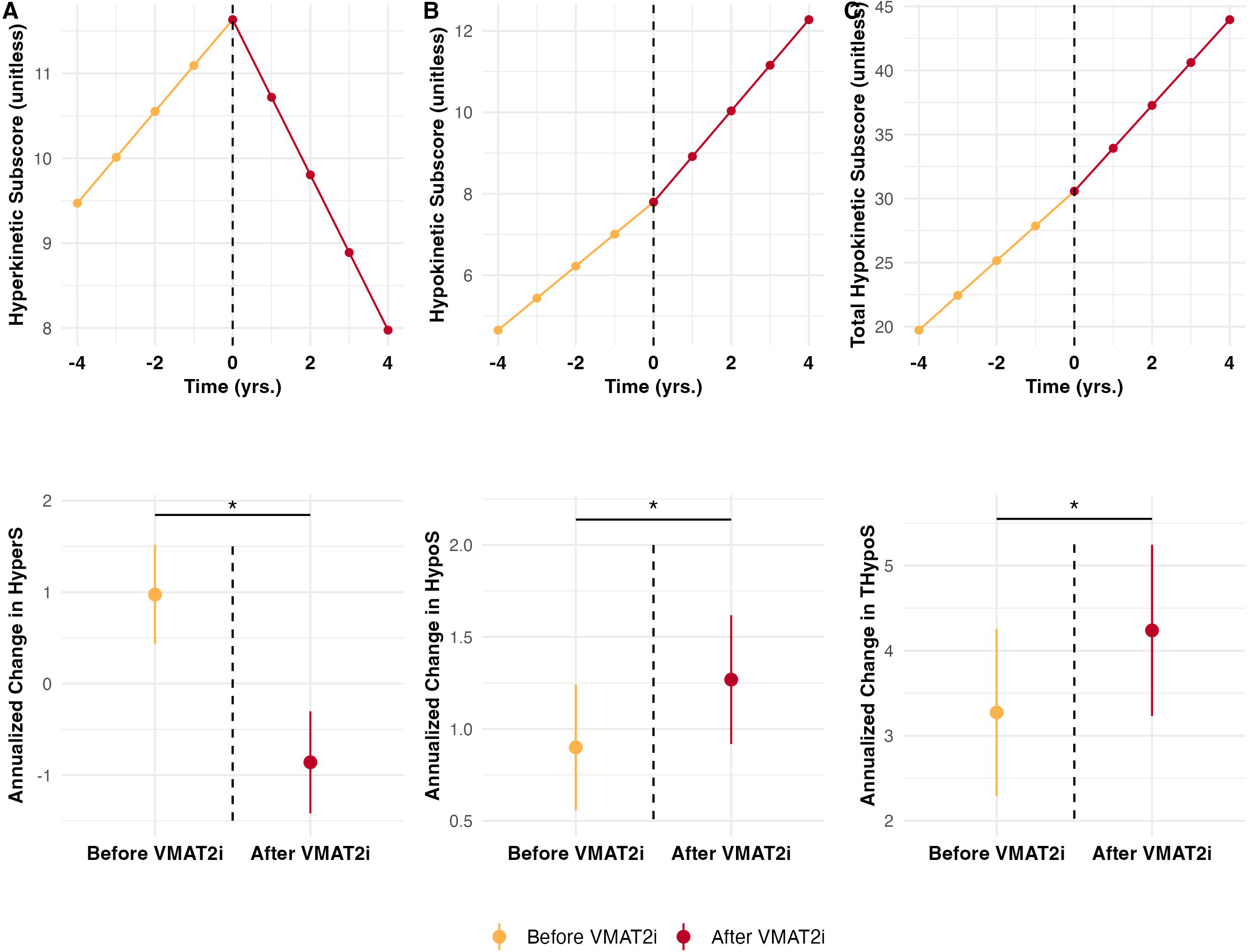
Plots of variation in rates of change in motor scores before and after VMAT2i administration. VMAT2i: vesicular monoamine transporter type-2 inhibitor. A bracket and asterisk denote a significant difference between slopes. Error bars denote standard error, not confidence intervals.

**Table 2:**
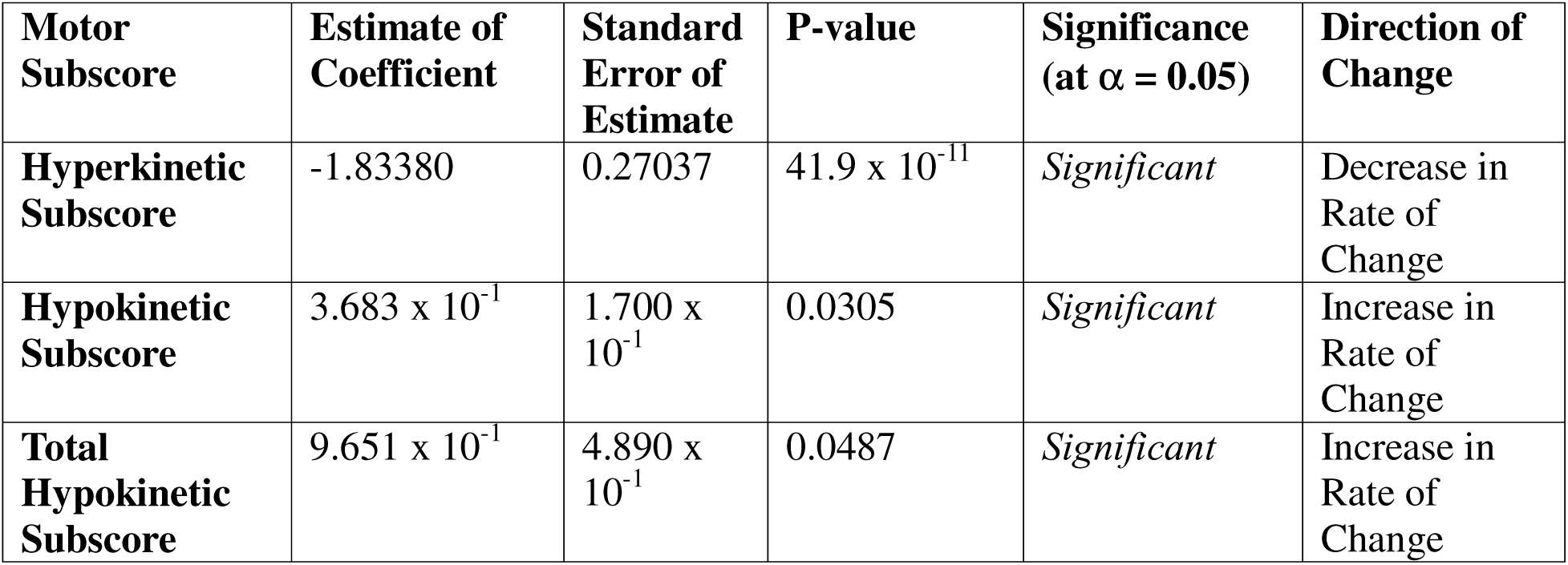
Results of VMAT2i analysis. VMAT2i: vesicular monoamine transporter type-2.

| **Motor Subscore** | **Estimate of Coefficient** | **Standard Error of Estimate** | **P-value** | **Significance (at $\alpha = 0.05$)** | **Direction of Change** |
| --- | --- | --- | --- | --- | --- |
| **Hyperkinetic Subscore** | -1.83380 | 0.27037 | $41.9 \times 10^{-11}$ | *Significant* | Decrease in Rate of Change |
| **Hypokinetic Subscore** | $3.683 \times 10^{-1}$ | $1.700 \times 10^{-1}$ | 0.0305 | *Significant* | Increase in Rate of Change |
| **Total Hypokinetic Subscore** | $9.651 \times 10^{-1}$ | $4.890 \times 10^{-1}$ | 0.0487 | *Significant* | Increase in Rate of Change |

Baseline demographics of the antipsychotic users are shown in *Supplementary Table 1.* The results of the antipsychotic analysis are found in *Supplementary Table 2*. Similar to VMAT2i’s, the mean annualized change in HyperS prior to antipsychotic initiation was 1.19 p.p.y. (S.E. 0.24), significantly decreasing to -0.12 p.p.y. (S.E. 0.24) after antipsychotic use (p < 0.0001; *Supplementary Figure 2A*). In the case of antipsychotics, neither the mean annualized change in HypoS (pre-antiψ: 1.34 ppy [S.E. 0.17], post-antiψ: 1.47 ppy [S.E. 0.17]) nor THypoS (pre-antiψ: 4.40 ppy [S.E. 0.44], post-antiψ: 4.52 ppy [S.E. 0.44]) were significantly different after administration. (p_HypoS_ = 0.322, p_THypoS_ = 0.742, *Supplementary Figure 2B-C*).

This information can be succinctly summarized by the mean differences between the annualized rates of change before / after starting the drugs of interest. In the case of VMAT2i’s, the mean differences are: -1.83 (95% C.I.: -2.36, -1.31) for HyperS, 0.368 (95% C.I.: 0.0360, 0.701) for HypoS and 0.965 (95% C.I.: 0.00919, 1.92) for THypoS. In the case of antipsychotics, the mean differences are: -1.31 (95% C.I.: -1.69, -0.921) for HyperS, 0.132 (95% C.I.: -0.129, 0.395) for HypoS and 0.118 (95% C.I.: -0.583, 0.819) for THypoS. The mean differences correspond to the p-values reported in the previous two paragraphs, with significant p-values indicating a 95% confidence interval that does not contain 0, and the reverse being true.

## Discussion

In this analysis we have shown that VMAT2i’s were associated with significant worsening of (classical and total) hypokinetic features of HD. VMAT2i’s were associated with a significant improvement in hyperkinesis, providing internal validity to this study. Importantly, the rates of worsening (*Δ HyperS_pre-VMAT2i_*= 0.97 p.p.y., S.E. 0.28) and improvement (*Δ HyperS_post-VMAT2i_*= -0.86 p.p.y., S.E. 0.28) in hyperkinesis were modest. However, this finding is in accordance with our previous work, which demonstrated the limited progression of hyperkinetic features. On average, individual hyperkinetic subscales only reach a peak severity of ∼37.5% (i.e., 1.5/4) (12). Nevertheless, this chorea, despite its ostensibly low rate of progression, can cause clinically significant dysfunction in patients (who may or may not possess insight into the matter) and role strain in caregivers. We report that the mean slope of worsening of hyperkinetic features was 0.97 p.p.y., which likely represents a clinically significant change. Consequently, reversing that slope to -0.86 p.p.y. with the use of VMAT2i’s reasonably represents a statistically and clinically significant improvement in hyperkinetic features. Furthermore, these slopes are model-derived and hence represent an aggregate of pooled data, which naturally will not apply to every patient in the same manner. Calculating the ‘raw’ annualized rate of change in HyperS prior to VMAT2i administration using the unmodeled data revealed a worsening of 1.2 p.p.y. (and 1.08 p.p.y. prior to antipsychotic administration), which are similar to the model-derived estimates.

The effect of VMAT2i’s on HypoS and THypoS was consistent with expectations; VMAT2i’s significantly worsened the annualized rate of change in both measures of hypokinesis. Several conclusions can be drawn from these results. Firstly, they seem to complement our previous study which categorized non-choreiform motor features of HD as hypokinetic. Importantly, this classification was based on subscore trajectory and not neuropathology.(12) Hence, external validation of this conclusion using drug data was warranted. If the alternative hypothesis were true (*i.e.*, if trajectory did not track pathoetiology and the motor features were actually hyperkinetic), we should observe a maintained or attenuated rate of progression in these subscales. Rather, we observe a worsening in their rate of progression, derived using a model that controls for the effects of improper matching (via single-subject design) and normal time-related disease progression. Indeed, *most* subscales were affected by VMAT2 inhibition, rather than just a few classically hypokinetic measures. VMAT2i’s effect on HypoS was quite plausible, given that the score is constituted of known hypokinetic features (i.e. rigidity, postural instability, bradykinesia). However, when more ambiguous features were added to the composite measure (*i.e.* THypoS), the rate of change still displayed a significant increase. As a result, the effect of VMAT2i’s extends not only to classical hypokinetic features but rather to all non-choreiform measures.

Notably, antipsychotics displayed different pharmacologic properties than VMAT2i’s, with regards to the measured outcomes. The mean difference in HyperS after VMAT2i’s was - 1.83, whereas this difference was -1.31 after antipsychotics. This difference in effect size, though modest, points to a stronger antidopaminergic effect with VMAT2i’s when compared to antipsychotics. The mean differences in HypoS and THypoS after VMAT2i’s were 0.368 and 0.965, and these differences were significant. Meanwhile, the mean differences in HypoS and THypoS after antipsychotics were 0.132 and 0.118, neither reaching significance. This demonstrates that VMAT2i’s, when prescribed at standard doses, possess a statistically and clinically significant effect of worsening hypokinesis that antipsychotics lack. This is potentially related to the fact that antipsychotics are often prescribed in HD for irritability rather than psychosis, hence the utilization of lower doses (see *Results*) with lesser potential for extrapyramidal side-effects.(25) In light of these findings, clinical decision-making regarding the use of VMAT2i’s and/or antipsychotics should involve careful consideration of the balance in severity between hyperkinetic and hypokinetic features in the patient.

Critically, this study was not designed to directly compare antipsychotics with VMAT2i’s. Although we can compare efficacy in chorea dampening between VMAT2i’s (*Δ HyperS*_post-VMAT2i_ = -0.86 p.p.y., S.E. 0.28) and antipsychotics (*Δ HyperS*_post-antiψ_ = -0.12 p.p.y., S.E. 0.24), it must be prefaced that much heterogeneity exists in the dosing regiments (loading and maintenance) of the two drug classes. Furthermore the two populations are distinct; psychiatric manifestations occur earlier in the disease course, which is when antipsychotics are most likely to be prescribed.(1, 25) Hence, it is argued that the populations of VMAT2i users and antipsychotic users were at different stages of the disease when they were initiated on their medications-of-interest, and may thus react disparately. Consulting the baseline characteristics of VMAT2i (*Table 1*) and antipsychotic users (*Supplementary Table 1*), we may directly compare the two populations for relevant disease markers. Through calculation of *t* / *χ*^2^ values, the degree of similarity between the two groups can be deduced. For age, AGAB, age at clinical diagnosis, age at death, CAG repeats, HD-ISS stage, disease duration and disease burden, the p-values were all non-significant (*p* = 0.672, 0.495, 0.938, 0.966, 0.689, 0.976, 0.147 and 0.310, respectively).

Where they did differ, however, is their distribution of baseline motor scores. VMAT2i users overall had significantly higher mean baseline scores for TMS, HyperS, HypoS and THypoS. These differences are most likely because psychiatric manifestations occur before motor impairment, hence leading to lower motor scores in patients at an earlier phase of the disease. Nevertheless, the aim of this study was not a one-to-one comparison of the two groups. Rather, the single-subject experimental design has the advantage of matching each drug-user to themselves, eliminating the need for similarity between the two populations.

There are several strengths of this study. The use of a large number of participants allowed for a well-powered investigation. The construction of linear mixed-effects models and use of single-subject design bolstered our analyses by nullifying the effects of inadequate matching, different baseline disease severities and differential rates of disease progression. To our knowledge, this study is the first of its kind to study the effects of VMAT2i’s on the entirety of the UHDRS motor battery. This demonstrated that VMAT2i use was associated with worsening of non-choreiform features of HD. As VMAT2i’s remain the only FDA-approved medications for the treatment of HD, they have garnered a reputation as the characteristic drugs of choice, used to improve both TMS and the motor phenotype in general.(26) Nevertheless, clinicians worldwide have made anecdotal observations that VMAT2i’s tend to make their patients more hypokinetic.(27) Here, the results of *Enroll-HD* were leveraged to demonstrate this observation in a statistically significant manner. Tetrabenazine and its derivatives remain excellent at dampening chorea, an effect which antipsychotics (via dopamine receptor blockade) also possess. However, it is in considering the effect on non-choreiform features that the two drug classes differ, and hence their utility in treating different HD phenotypes as well. Therefore, these results may lead to more personalized and optimized care for HD patients.

Despite these strengths, there are several limitations of this work. First and foremost, this was an observational study. As a result, these findings represent associations and cannot infer causality. Second, direct comparison between different drugs classes is limited due to structural aspects of the *Enroll* dataset. Therefore, observed differences between the effects of antipsychotics and VMAT2i’s are just that, observations. Third, data on the indications for VMAT2i prescriptions is lacking. The reasons why HD patients are prescribed antipsychotics versus VMAT2i’s are certainly different, implying an underlying constitutional difference between the two groups that has yet to be elucidated. Future studies may include a superior experimental design to avoid these drawbacks. With regards to study design, participants were selected only if they continually took a VMAT2i up to and including the next visit, making undetected adverse events less likely. However, because most VMAT2i trials began sometime during the approximately one-year inter-visit interval, the immediate effects of the drug are unknowable. This is especially relevant given the drug’s hyperacute onset of action.(28) Hence, it is possible that more participants faced acute hypokinetic reactions hours-to-days after drug administration than was recorded at the 1-year follow-up. Furthermore, most participants possessed only the minimum three visit day structure, limiting conclusions on the long-term effects of continual VMAT2 inhibition on hyperkinetic and hypokinetic subscale progression.

## Data Availability

The datasets utilized for this secondary analysis are obtainable from the Enroll-HD clinical research platform, courtesy of CHDI, for use by professional researchers in their studies on HD.

https://enroll-hd.org/for-researchers/access-data-biosamples/

## Acknowledgements

This research is made possible due to the efforts of CHDI, the *Enroll-HD* clinical research platform and the participation of HD patients and their families.

## Authors’ Roles

1. Research Project: A. Conception, B. Organization, C. Execution; 2. Statistical Analysis: A. Design, B. Execution, C. Review and Critique; 3. Manuscript Preparation: A. Writing the First Draft, B. Review and Critique.

N.M.H.: 1A, 1B, 1C, 2A, 2B, 3A, 3B

P.C.N.: 3B

A.K.: 3B

## Disclosures

### Ethical Compliance Statement

We acknowledge having reviewed the Journal’s stance on ethical publication and confirm that our work adheres to these guidelines. The authors state that the data used in this secondary analysis was obtained from the studies *Enroll-HD* and *Registry*, which were conducted in accordance with local ethics board approval. The University of Iowa Institutional Review Board (IRB) waived approval for this paper, as the data utilized was de-identified, and participants’ consent for usage of their information in secondary analyses was obtained at the original study locations by CHDI.

## Financial disclosures from previous 12 months

The authors report no sources of funding and no conflicts of interest.

