## Supplement (Figures and Tables) for "Effect of Vesicular Monoamine Transporter Type-2 Inhibitors on Motor Features in HD"

*Supplementary Figure 1*

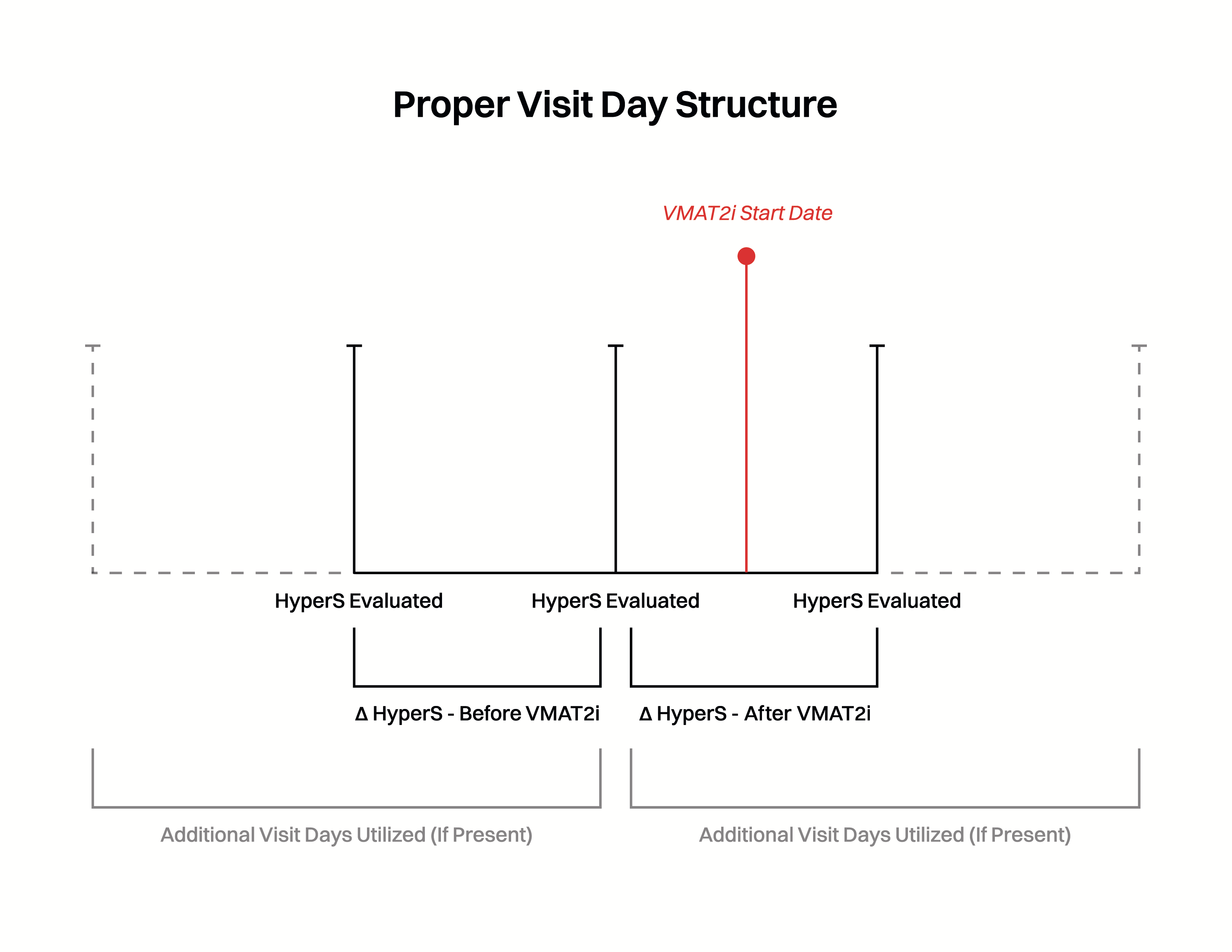

*Supplementary Figure 2*

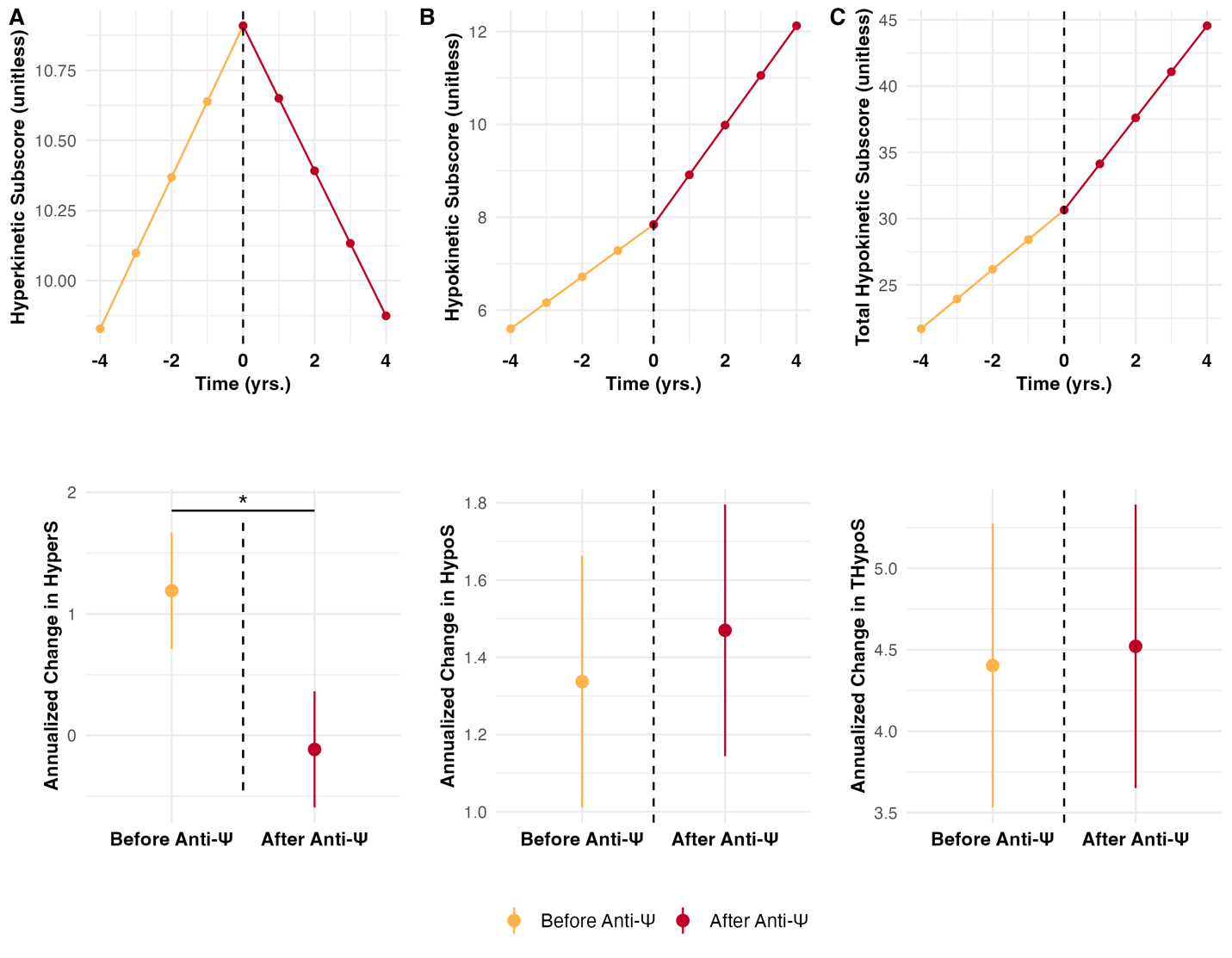

*Supplementary Table 1:*

| **Demographic** | **Value** |
| --- | --- |
| **Age (yrs.)** |  |
| Mean (SD) | 51.4 (11.9) |
| Median [Min, Max] | 51.5 [21.0, 84.0] |
| **Assigned Gender at Birth** |  |
| Assigned Male at Birth (%) | 484 (49.8%) |
| Assigned Female at Birth (%) | 488 (50.2%) |
| **Calculated BMI (kg/m^2^)** |  |
| Mean (SD) | 24.5 (4.77) |
| Median [Min, Max] | 23.7 [14.0, 62.8] |
| Missing (%) | 35 (3.6%) |
| **Duration of Disease (yrs.)** |  |
| Mean (SD) | 3.54 (3.79) |
| Median [Min, Max] | 2.00 [-2.00, 28.0] |
| Missing | 4 (0.4%) |
| **Age at Clinical Diagnosis** |  |
| Mean (SD) | 47.8 (11.7) |
| Median [Min, Max] | 48.0 [17.0, 83.0] |
| Missing (%) | 4 (0.4%) |
| **Use of VMAT2i** |  |
| Users (%) | 248 (25.5%) |
| Never-Users (%) | 724 (74.5%) |
| **Disease Burden** |  |
| Mean (SD) | 413 (88.3) |
| Median [Min, Max] | 408 [108, 918] |
| **Age of Death (yrs.)** |  |
| Mean (SD) | 58.7 (11.8) |
| Median [Min, Max] | 58.0 [29.0, 85.0] |
| Missing (%) | 846 (87.0%) |
| **Cause of Death** |  |
| Pneumonia (%) | 29 (3.0%) |
| Other Infection (%) | 5 (0.5%) |
| Cancer (%) | 6 (0.6%) |
| Stroke (%) | 0 (0.0%) |
| Trauma (%) | 1 (0.1%) |
| Suicide (%) | 8 (0.8%) |
| Other (%) | 46 (4.7%) |
| Missing (%) | 877 (90.2%) |
| **Total Motor Score** |  |
| Mean (SD) | 33.1 (6.5) |
| Median [Min, Max] | 31.0 [0, 105] |
| **Hyperkinetic Subscore (HyperS)** |  |
| Mean (SD) | 9.17 (4.71) |
| Median [Min, Max] | 9.00 [0, 26.0) |
| **Hypokinetic Subscore (HypoS)** |  |
| Mean (SD) | 5.97 (4.15) |
| Median [Min, Max] | 5.00 [0, 26.0] |
| **Total Hypokinetic Subscore (THypoS)** |  |
| Mean (SD) | 23.9 (14.1) |
| Median [Min, Max] | 21.0 [0, 86.0] |
| **Ethnicity** |  |
| White (%) | 918 (94.4%) |
| Black (%) | 5 (0.5%) |
| Latinx (%) | 16 (1.6%) |
| Native American (%) | 1 (0.1%) |
| Mixed (%) | 6 (0.6%) |
| Asian (%) | 7 (0.7%) |
| Other (%) | 19 (2.0%) |
| **CAG Repeats** |  |
| Mean (SD) | 44.2 (3.55) |
| Median [Min, Max] | 43.0 [37.0, 67.0] |
| **HD-ISS Stage** |  |
| 0 | 0 (0%) |
| 1 | 2 (0.2%) |
| 2 | 58 (6.0%) |
| 3 | 517 (53.2%) |
| Missing | 395 (40.6%) |
| **Average Antipsychotic Total Daily Dose by Three Most Common Medications (in milligrams, 3 significant figures)** |  |
| Olanzapine (most common) | 5.84 |
| Risperidone (second-most common) | 1.54 |
| Quetiapine (third-most common) | 81.1 |

*Supplementary Table 2:*

| **Motor Subscore** | **Estimate of Coefficient** | **Standard Error of Estimate** | **P-value** | **Significance (at α = 0.05)** | **Direction of Change** |
| --- | --- | --- | --- | --- | --- |
| **Hyperkinetic Subscore** | -1.30645 | 0.19680 | 4.11 x 10^-11^ | *Significant* | Decrease in Rate of Change |
| **Hypokinetic Subscore** | 0.13286 | 0.13406 | 0.32181 | Non-Significant | Increase in Rate of Change |
| **Total Hypokinetic Subscore** | 0.11797 | 0.35821 | 0.74194 | Non-significant | Increase in Rate of Change |

**Legends:**

*Supplementary Figure 1:* Structure of study design. Δ HyperS: annualized change in hyperkinetic subscore; VMAT2i: vesicular monoamine transporter type-2 inhibitor.

*Supplemental Figure 2:* Plots of variation in rates of change in motor scores before and after antipsychotic medication administration. A bracket and asterisk denote a significant difference between slopes. Error bars denote standard error, not confidence intervals.

*Supplemental Table 1:* Baseline demographics of antipsychotic medications users. Yrs: years; BMI: body mass index; kg: kilogram; m^2^: squared meter; SD: standard deviation; VMAT2i: vesicular monoamine transporter type-2 inhibitors; CAG: cytosine-adenine-guanine; min: minimum; max: maximum.

*Supplemental Table 2:* Results of antipsychotic medications analysis.
